# Targeting MAdCAM-1 can prevent colitic cancer progression by suppressing immune cell infiltration and inflammatory signals

**DOI:** 10.1101/2022.12.18.22283633

**Authors:** Naoya Ozawa, Takehiko Yokobori, Katsuya Osone, Erkhem-Ochir Bilguun, Haruka Okami, Yuki Shimoda, Takuya Shiraishi, Takuhisa Okada, Akihiko Sano, Makoto Sakai, Makoto Sohda, Tatsuya Miyazaki, Munenori Ide, Hiroomi Ogawa, Takashi Yao, Tetunari Oyama, Ken Shirabe, Hiroshi Saeki

## Abstract

Chronic inflammation by infiltrating immune cells promotes colitis-associated dysplasia/colitic cancer in ulcerative colitis (UC) via activating inflammatory cytokine signalling (IL-6/p-STAT3 and TNFα/NF-κB). Mucosal addressin cell adhesion molecule-1 (MAdCAM-1) is a cell adhesion molecule expressed on high endothelial venules that promote immune cell migration from the bloodstream to the gut. MAdCAM-1 targeting strategy is attracting attention as a novel therapeutic option for UC. However, the significance of MAdCAM-1-positive vessels in dysplasia/colitic cancers remains unclear. We conducted immunohistochemistry against MAdCAM-1, and immune cell markers in surgically resected samples from 11 UC patients with dysplasia/colitic cancer and 17 patients with sporadic colorectal cancer (SCRC). Moreover, we used a colitic cancer model, azoxymethane (AOM)/dextran sodium sulphate (DSS) mouse, to evaluate whether anti-MAdCAM-1 blocking antibody can suppress colitic cancer progression. MAdCAM-1-positive vessel number and infiltrating CD8-, CD68-, and CD163-positive immune cell numbers were significantly higher in dysplasia/colitic cancer than in normal mucosa, SCRC, and UC mucosa. In the AOM/DSS mouse model, MAdCAM-1 antibody reduced the tumour number, tumour diameter, number of CD8-, CD68-, and CD163-positive immune cells, and IL-6/p-STAT3 and TNFα/NF-κB expression levels. Targeting MAdCAM-1 could be promising for inflammatory carcinogenesis, and tumour progression by regulating inflammation/immune cell infiltration in patients with UC.

**Lay summary:** MAdCAM-1 targeting strategy can control ulcerative colitis severity, carcinogenesis, and tumour progression by regulating inflammation/immune cell infiltration in patients with ulcerative colitis.

## Introduction

Chronic inflammation has been suggested to cause colorectal carcinogenesis in sporadic colorectal cancer (SCRC), and anti-inflammatory drugs have been reported to prevent such phenomena (1). In contrast, autoimmune responses by infiltrating immune cells can promote chronic inflammation and rare colitis-associated dysplasia/colitic cancer in the colorectal mucosa of patients with ulcerative colitis (UC) via activation of inflammatory cytokines (IL-6 and TNFα) and tumour-promoting transcription factors such as p-STAT3 and NF-κB (2, 3). Therefore, the blockade of immune cell migration into the UC mucosa by anti-inflammatory drugs has been a promising therapeutic strategy to reduce inflammatory disease activity in patients with UC (4). However, it is controversial whether blocking the infiltration of immune cells into the UC mucosa can prevent the initiation and progression of inflammatory dysplasia/colitic cancer or facilitate colorectal tumorigenesis via suppression of tumour immunity (5, 6).

Mucosal addressin cell adhesion molecule-1 (MAdCAM-1) is a cell adhesion molecule expressed in high endothelial venules (HEVs) in gut-associated lymphoid tissues and mesenteric lymph nodes (7). The binding of α4β7 integrin on immune cells to MAdCAM-1-positive vessels promotes immune cell migration from the bloodstream to intestinal lymphoid tissues. Interestingly, MAdCAM-1-positive vessels are strongly induced in HEV-like vessels of chronic inflammatory intestinal mucosa caused by inflammatory bowel diseases (IBDs) such as UC and Crohn’s disease (8), promoting the infiltration of α4β7 integrin-positive immune cells and contributing to disease progression in IBD (9).

The interaction between α4β7 integrin on immune cells and MAdCAM-1 on the vascular endothelium functions in an intestinal-specific manner and is a promising therapeutic target to suppress immune cell infiltration into the inflammatory colorectal mucosa of patients with UC (10). Vedolizumab, an α4β7 integrin inhibitor, has been used successfully in patients with UC. In addition, the MAdCAM-1 targeting strategy, which is the focus of this study, is also attracting attention as a novel therapeutic option for refractory UC via the direct inhibition of inflammatory immune cell infiltration (11). Therefore, a phase III clinical trial to evaluate the therapeutic efficacy of the MAdCAM-1 antibody in patients is currently ongoing (12). However, to date, the significance of MAdCAM-1-positive vessels on immune cell infiltration in clinical dysplasia/colitic cancer samples and the potential of MAdCAM-1-targeting in animal models have not been investigated.

This study aimed to clarify the importance of MAdCAM-1-positive vessels to infiltrating immune cells in the colorectal mucosa of patients with dysplasia/colitic cancer and SCRC. Therefore, we performed immunohistochemical staining for MAdCAM-1 and immune cell markers, such as CD8 (cytotoxic T cell marker), CD68 (macrophage marker), CD163 (tumour-associated macrophage marker), and FOXP3 (regulatory T cell marker), in surgically resected samples from 17 SCRC patients and 11 UC patients with colorectal dysplasia/colitic cancer. Moreover, we conducted an animal experiment as a colitic cancer model to evaluate whether the MAdCAM-1 antibody as an anti-inflammatory treatment can suppress dysplasia/colitic cancer progression via the regulation of immune cell infiltration into the colorectal mucosa. Our results demonstrate the promising potential of the MAdCAM-1 targeting strategy to prevent tumour aggressiveness and inflammatory immune cell infiltration in UC patients with rare dysplasia/colitic cancer.

## Results

### Clinical significance of MAdCAM-1-positive vessels in tissues of patients with SCRC and dysplasia/colitic cancer

We performed immunohistochemistry to detect a chronic inflammation-induced increase in MAdCAM-1-positive and whole intestinal vessels with CD31 expression in the clinical specimens of patients with SCRC and dysplasia/colitic cancer (Figure 1A). MAdCAM-1-positive vessels in UC were higher than that in normal mucosa, and those in dysplasia and colitic cancer were higher than that in the normal mucosa, SCRC, and UC (Figure 1B). The MAdCAM-1-/CD31-positive vessel ratio in dysplasia and colitic cancer was higher than that in normal mucosa, SCRC (Figure 1C), suggesting a specific increase in chronic inflammation-induced MAdCAM-1-positive vessels in UC and dysplasia/colitic cancer, rather than an increase in vessel number as a whole.

**Figure 1.**
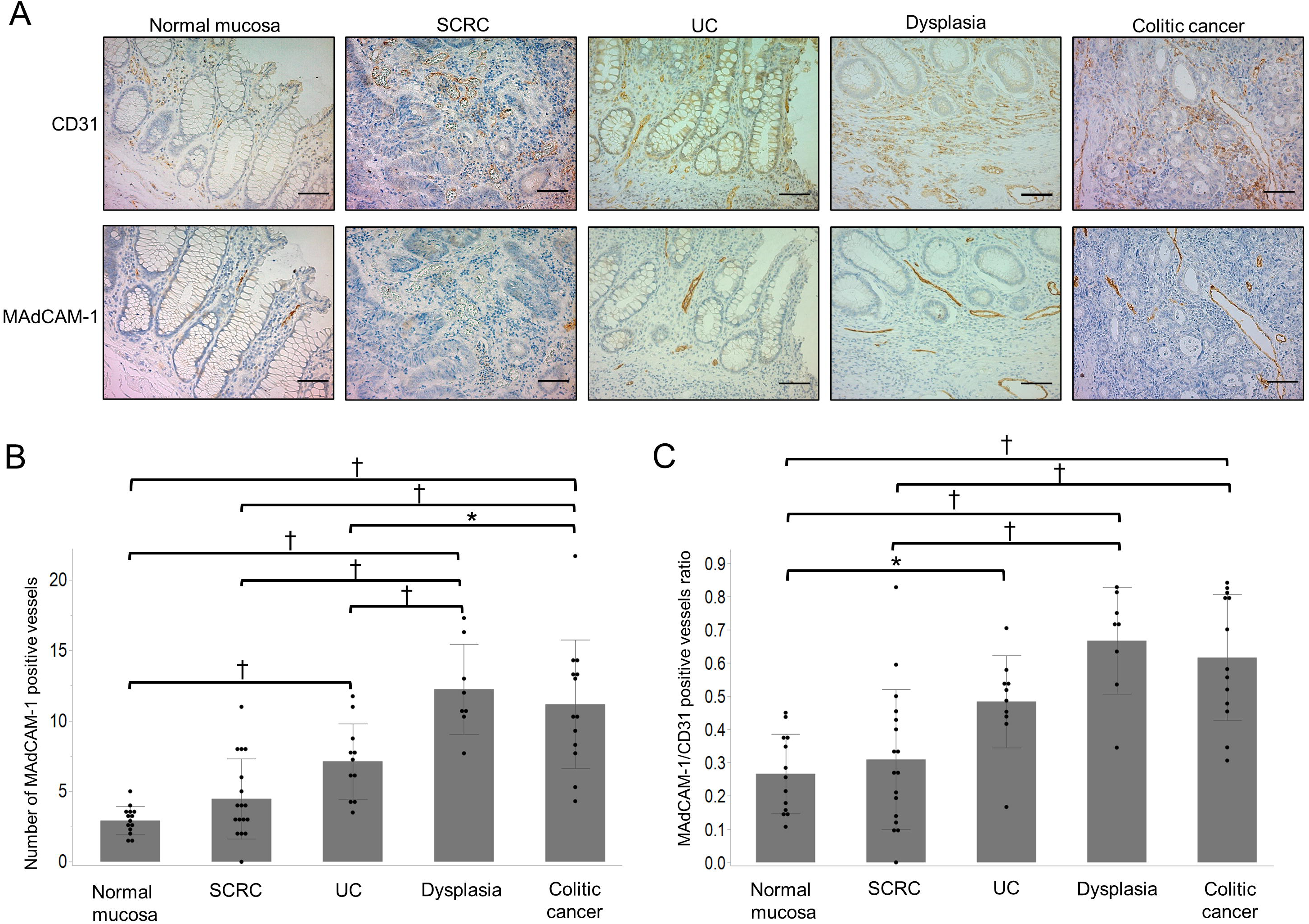
Immunohistochemical staining of CD31 and MAdCAM-1 in normal, SCRC, UC, dysplasia, and colitic cancer mucosa. (A) Upper panel shows the CD31- positive vessels in normal, SCRC, UC, dysplasia, and colitic cancer mucosa. The lower panel shows the MAdCAM-1-positive vessels in normal, SCRC, UC, dysplasia, and colitic cancer mucosa. Scale bar, 100 μm. (Original magnification, ×200). (B) The number of MAdCAM-1-positive vessels in normal mucosa (n=14), SCRC (n=17), UC (n=11), dysplasia (n=8), and colitic cancer (n=13). (C) The ratio of MAdCAM-1-positive vessels to CD31-positive vessels in normal (n=14), SCRC (n=17), UC (n=11), dysplasia (n=8), and colitic cancer (n=13) mucosa. These data are expressed as the mean ± SD. * P<0.05, †P<0.01. SCRC, sporadic colorectal cancer; UC, ulcerative colitis; MAdCAM-1, mucosal addressin cell adhesion molecule-1.

### Correlation between MAdCAM-1-positive vessels and immune cell infiltration in tissues of patients with SCRC and dysplasia/colitic cancer

Immunohistochemistry was performed to evaluate the relation between MAdCAM-1- positive vessels and immune cell markers, such as CD8 (cytotoxic T cell marker), CD68 (macrophage marker), CD163 (tumour-associated macrophage marker), and FOXP3 (regulatory T cell marker), in the clinical specimens of patients with SCRC and dysplasia/colitic cancer (Figure 2A). The number of infiltrating immune cells with CD8, CD68, and CD163 expression were higher in dysplasia/colitic cancer than in SCRC (Figure 2B). We divided the 21 dysplasia/colitic cancer tissues into low MAdCAM-1 (n=10) and high MAdCAM-1 (n=11) groups according to the MAdCAM-1-positive vessel number. Supplemental Table 1 and Figure 3 show the relationship between MAdCAM-1-positive vessel number, clinicopathological factors, and immune cell infiltration in dysplasia/colitic cancer samples. The high MAdCAM-1 group was associated with high infiltration of CD8- and CD163-positive immune cells; however, no significant differences were observed in age, sex, tumour location and differentiation, T factor, N factor, M factor, or infiltration of CD68- and FOXP3-positive cells between the high MAdCAM-1 and low MAdCAM-1 groups (Figure 3, Supplemental Table 2).

**Figure 2.**
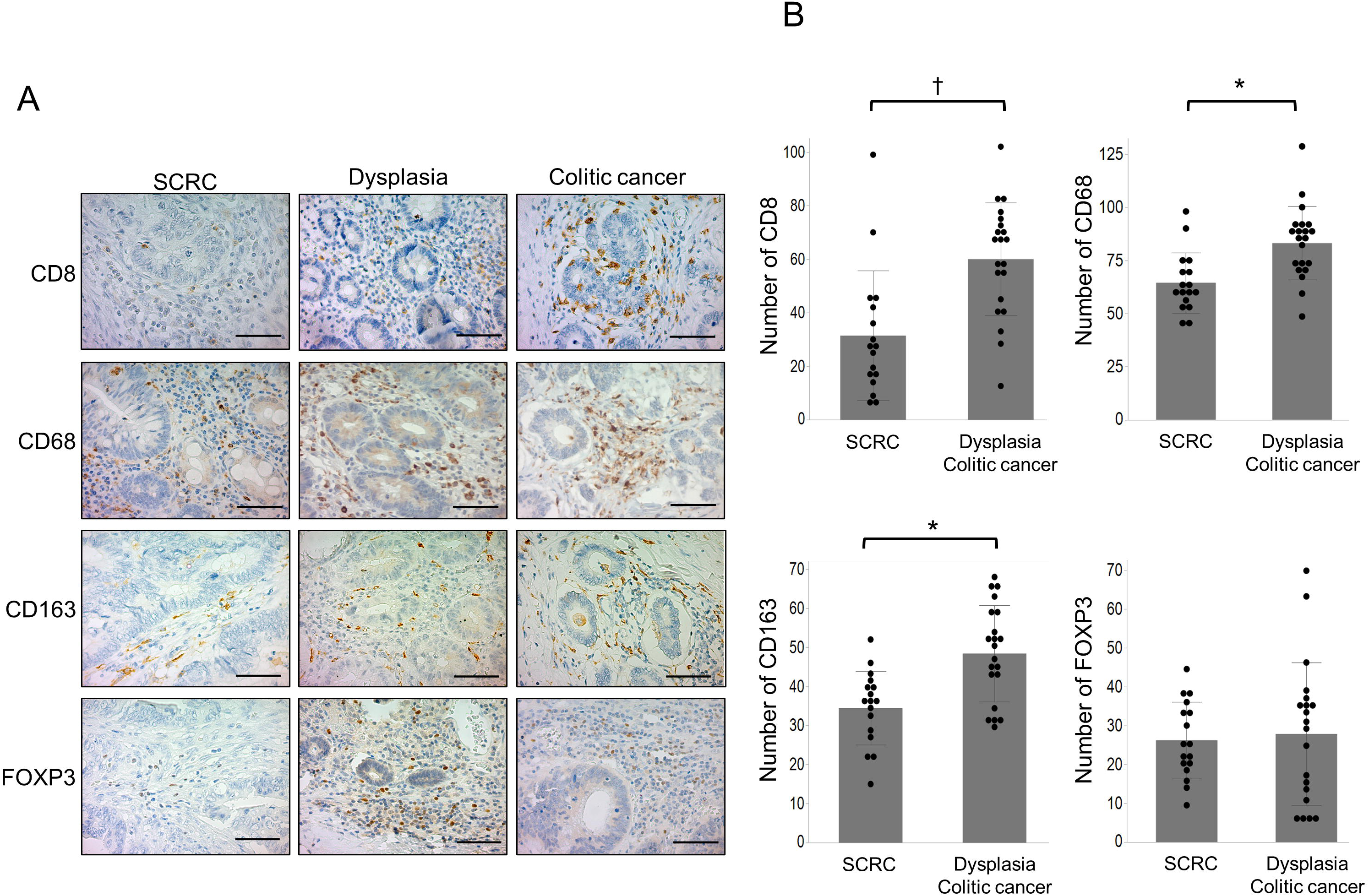
Immunohistochemical staining of CD8, CD68, CD163, and FOXP3 in SCRC, dysplasia, and colitic cancer tissues. (A) Representative immunohistochemical images of immune cell markers such as CD8 (cytotoxic T cell marker), CD68 (macrophage marker), CD163 (tumour-associated macrophage marker), and FOXP3 (regulatory T cell marker) in SCRC (n=17), dysplasia (n=8), and colitic cancer (n=13) tissues. Scale bar, 50 μm. (Original magnification, ×400). (B) The number of CD8-, CD68-, CD163-, and FOXP3-positive immune cells in SCRC (n=17), dysplasia (n=8), and colitic cancer (n=13). The data are expressed as the mean ± SD. *P<0.05, †P<0.01. SCRC, sporadic colorectal cancer.

**Figure 3.**
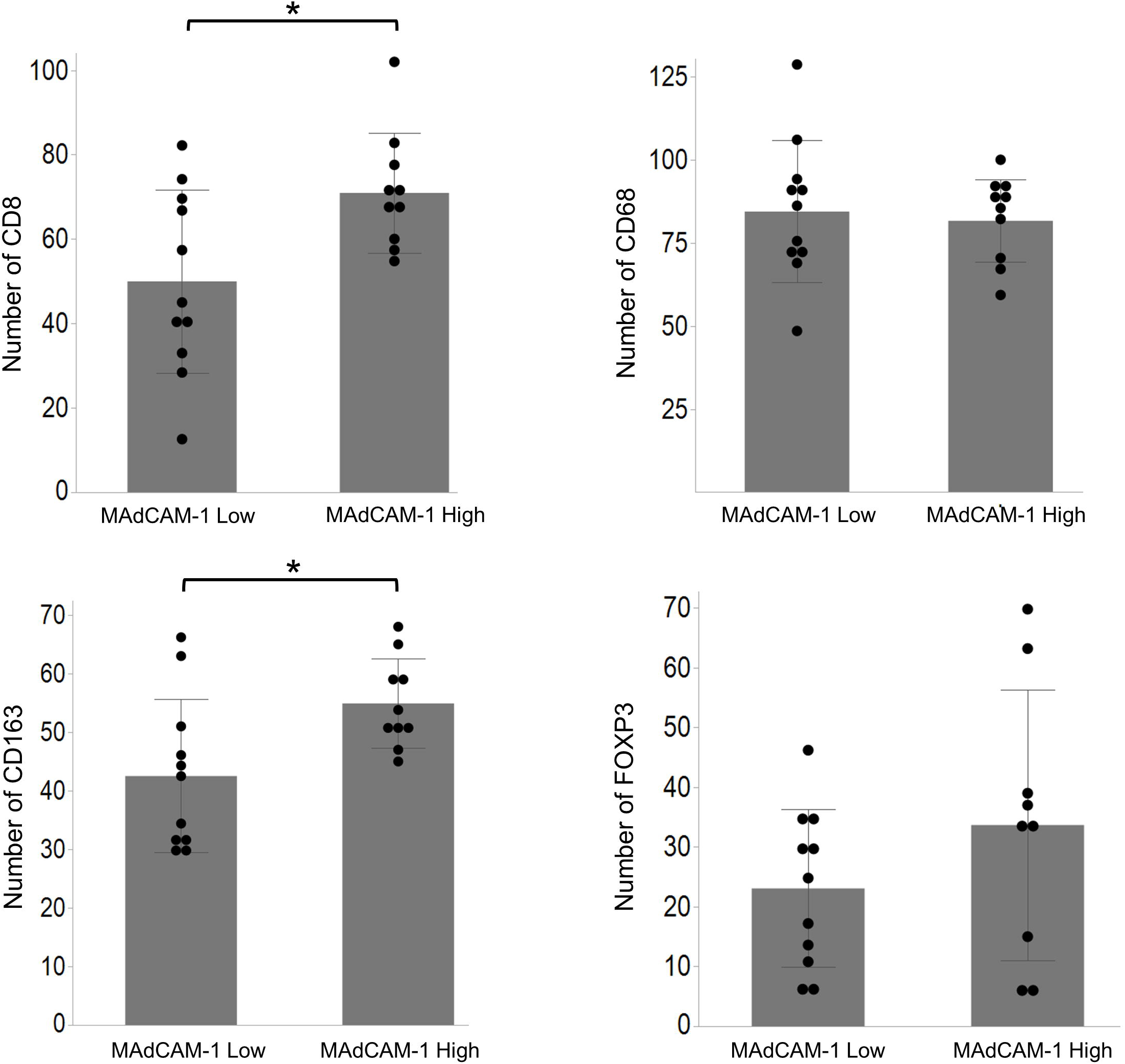
The relationship between the MAdCAM-1-positive vessels and immune cells in dysplasia/colitic cancer tissues. UC patients with dysplasia/colitic cancer tissues (n=21) were divided into low MAdCAM-1 (n=10) and high MAdCAM-1 (n=11) groups. The high MAdCAM-1 group had increased numbers of immune cells with CD8 and CD163 expression compared to the low group. These data are expressed as mean ± SD. *P<0.05. UC, ulcerative colitis; MAdCAM-1, mucosal addressin cell adhesion molecule-1.

### MAdCAM-1 antibody treatment improves colitis in an experimental colitic cancer mouse model

We investigated the therapeutic effect of the MAdCAM-1 blocking antibody in azoxymethane (AOM)/dextran sodium sulphate (DSS) mice, a representative experimental colitic cancer mouse model (Figure 4A). The disease activity index (DAI) score in the MAdCAM-1 Ab group was significantly lower than that in the IgG Ab group during all three DSS drinking periods (Figure 4A). Figure 4B shows a representative H&E-stained section of the colon mucosa of the mouse model, indicating high levels of immune cell infiltration in the IgG Ab group compared to that in the MAdCAM-1 Ab group (Figure 4B). In addition, the histological colitis score of the MAdCAM-1 Ab group was significantly lower than that of the IgG Ab group (Figure 4C).

**Figure 4.**
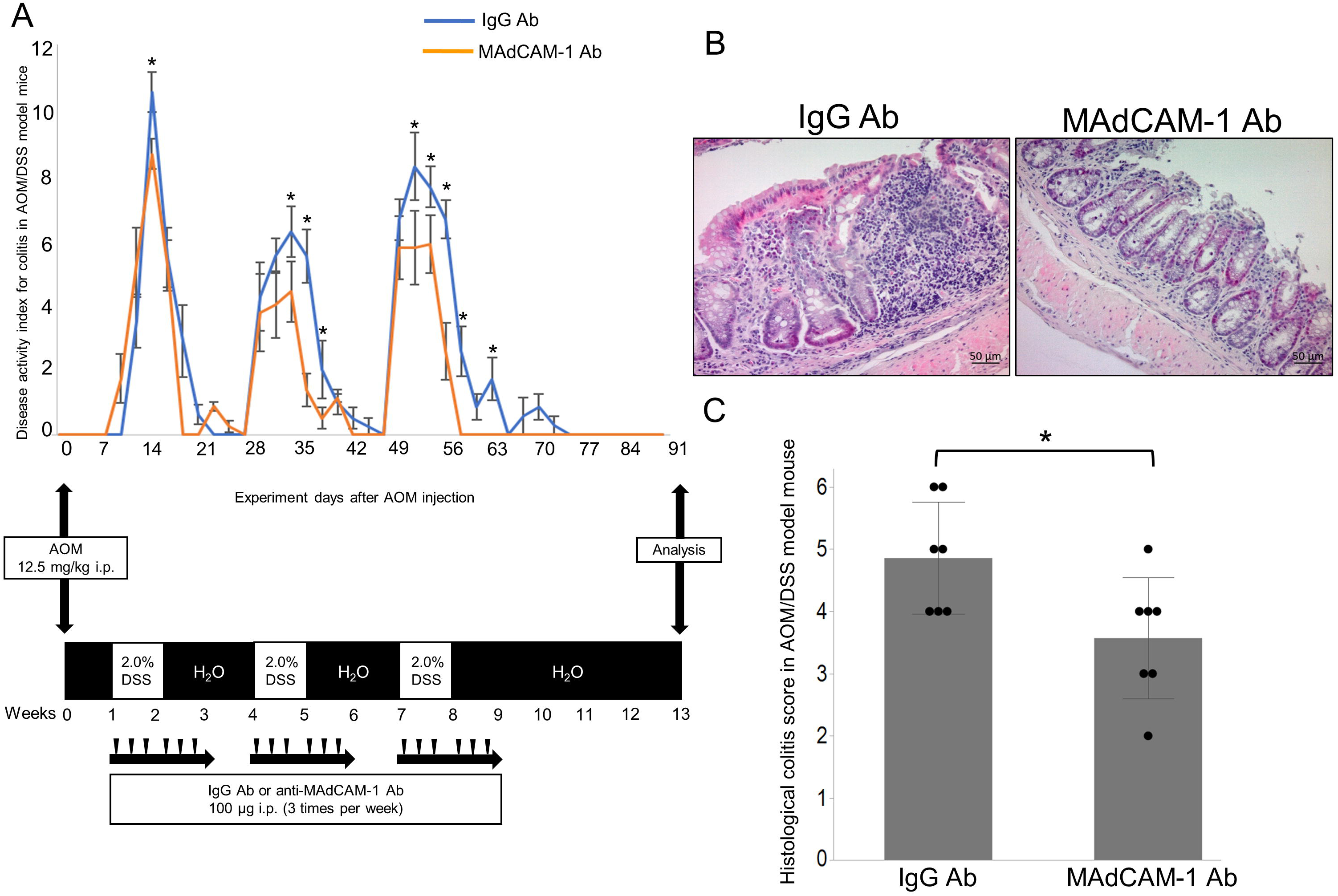
Therapeutic effect of MAdCAM-1 antibody in an experimental colitic cancer mouse model. (A) Upper graph shows the disease activity index score in the IgG Ab (n=7) and MAdCAM-1 Ab (n=7) groups of AOM/DSS mice experiments. The lower graph shows drug treatment schedules of AOM/DSS mice administered with 12.5 mg/kg AOM and 2.5% DSS as described in the Materials and methods section. (B) Representative H&E images of colon mucosa of the IgG Ab and the MAdCAM-1 Ab groups. (C) The histological colitis score in the IgG Ab (n=7) and the MAdCAM-1 Ab (n=7) groups. The data are expressed as the mean ± SD. *P<0.05. MAdCAM-1, mucosal addressin cell adhesion molecule-1; AOM, azoxymethane; DSS, dextran sodium sulphate.

### Anti-tumour effect of the MAdCAM-1 antibody in AOM/DSS mouse

To investigate the anti-tumour effects of MAdCAM-1 antibody, we measured the number and size (average diameter in millimetres) of tumours that developed in the colons harvested from the two groups (Figure 5A). The number and size of the tumours in the MAdCAM-1 Ab group were significantly lower than those in the IgG Ab group (Figure 5B). The number of cases of tumour invasion into the lamina propria mucosa in the MAdCAM-1Ab group was significantly lower than that in the IgG Ab group (Figure 5C). The percentage of the proliferation marker Ki67-positive cells in the tumours of the MAdCAM-1Ab group was significantly lower than that of the IgG Ab group (Figure 5D).

**Figure 5.**
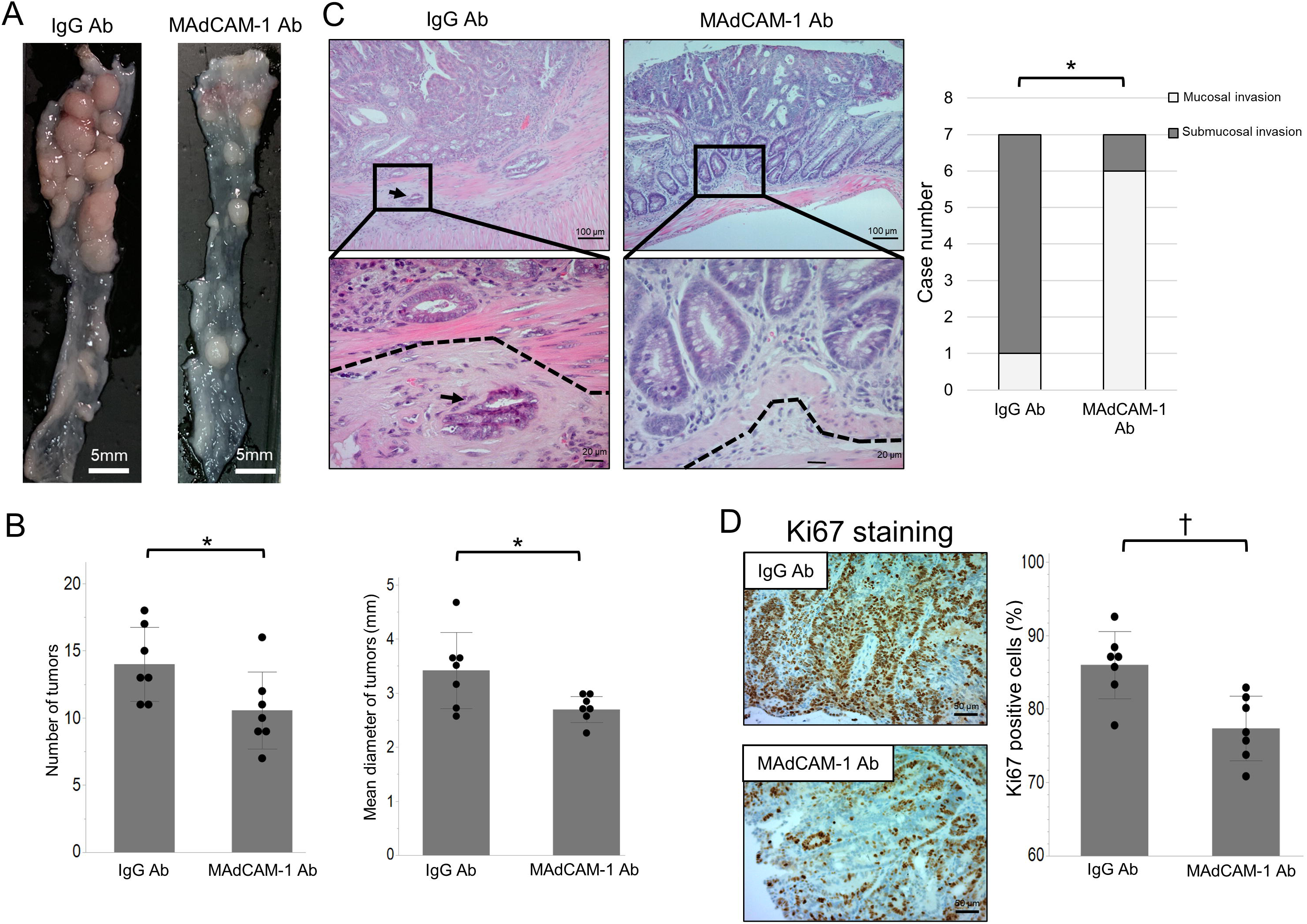
Impact of the MAdCAM-1 antibody treatment on the colon tumorigenesis and progression in AOM/DSS mouse model. (A) Representative macroscopic images of colon tumours in the IgG Ab and the MAdCAM-1 Ab groups of AOM/DSS mice. (B) Macroscopic tumour number and mean tumour diameters in the IgG Ab (n=7) and MAdCAM-1 Ab (n=7) groups. (C) Representative H&E sections of colon tumours in the IgG Ab and the MAdCAM-1 Ab groups (original magnification, ×200). Arrow indicates the submucosal invasion in the IgG Ab group. The dotted line indicates the border between the muscularis mucosa and submucosal layers. (D) Representative immunohistochemical staining images of Ki67 in the IgG Ab (n=7) and the MAdCAM-1 Ab (n=7) groups. The percentage of Ki67-positive tumour cells in the IgG Ab group was significantly higher than those in the MAdCAM-1 Ab group. The data are expressed as the mean ± SD. *P<0.05. MAdCAM-1, mucosal addressin cell adhesion molecule-1; AOM, azoxymethane; DSS, dextran sodium sulphate.

### Correlation between MAdCAM-1 antibody treatment and immune cell infiltration in the tumours of AOM/DSS mice

We performed immunohistochemistry to evaluate the infiltration levels of immune cells, such as CD8-, CD68-, CD163-, and FOXP3-positive cells, in the two groups of AOM/DSS mice (Figure 6A). The number of infiltrating immune cells with CD8, CD68, and CD163 expression was lower in the MAdCAM-1Ab group than in the IgG Ab group (Figure 6B). Furthermore, we evaluated the expression levels of IL-6, TNFα, perforin, IL-10, and TGF-β as representative cytokines from immune cells in the two groups using RT-PCR. The expression of IL-6 and TNFα in the MAdCAM-1 Ab group was significantly lower than that in the IgG Ab group (Figure 6C), suggesting that MAdCAM-1 antibody treatment suppressed the expression levels of cytokines such as IL-6 and TNFα via downregulation of the infiltration of T cells and macrophages into the tumours of the colitic cancer mouse model. There were no significant differences in the levels of other cytokines between the MAdCAM-1 Ab and IgG Ab groups (Supplemental Figure 1).

**Figure 6.**
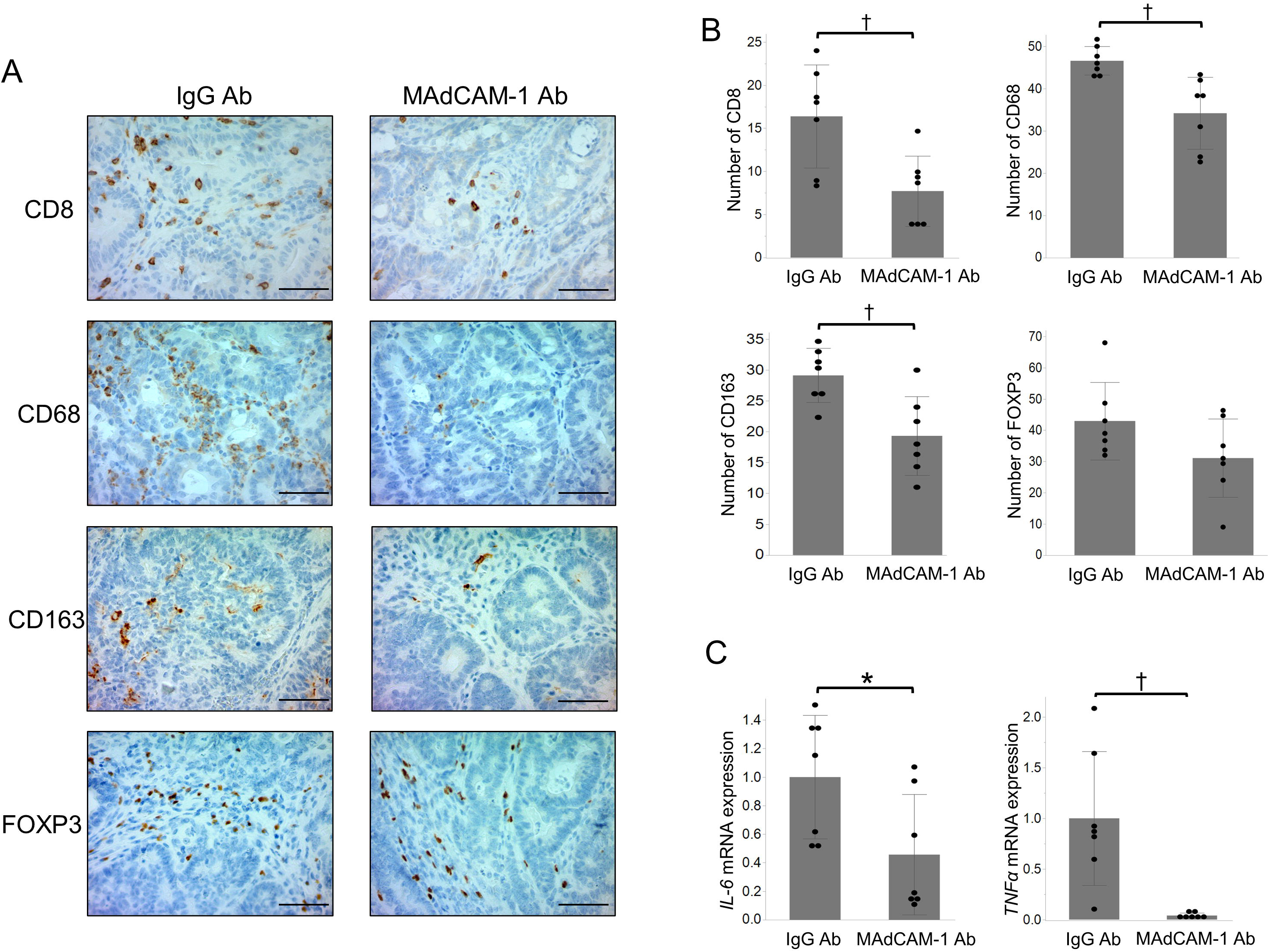
Significance of the MAdCAM-1 antibody treatment on immune cell infiltration and cytokine expression of colon tumours in the AOM/DSS mouse model. (A) Representative immunohistochemical images of CD8-, CD68-, CD163-, and FOXP3-positive cells in the colon tumours of IgG Ab and MAdCAM-1 Ab groups. Scale bar, 50 μm. (Original magnification, ×400). (B) CD8-, CD68-, CD163-, and FOXP3-positive cells in the colon tumours of IgG Ab (n=7) and MAdCAM-1 Ab (n=7) groups. (C) Relative expression levels of inflammatory cytokines *IL-6* and *TNFA* were examined in the tumours of the IgG Ab (n=7) and MAdCAM-1 Ab (n=7) groups. *GAPDH* was used as the internal reference in each group. The data are expressed as the mean ± SD. *P<0.05, †P<0.01. MAdCAM-1, mucosal addressin cell adhesion molecule-1.

### MAdCAM-1 antibody treatment inhibits the activation of p-STAT3 and NF-kB in the tumours of AOM/DSS mice

To clarify the molecular mechanism of the MAdCAM-1 antibody against tumours of the colitic cancer models, cap analysis of gene expression (CAGE) analysis was performed using tumours of the AOM/DSS mice from the IgG Ab and MAdCAM-1 Ab groups. Based on gene ontology (GO) analysis, the activity of cytokines, immune cells, angiogenesis, and proliferation was significantly downregulated in colitic tumours of the MAdCAM-1 Ab group compared to that in the IgG Ab group (Figure 7A). Furthermore, consistent with the GO analysis results, the expressions of several inflammation-related genes related to interleukin, TNFα, STAT, and NF-κB signalling were downregulated in the MAdCAM-1 Ab group compared to those in the IgG Ab group (Supplemental Figure 2). To validate the correlation between MAdCAM-1 antibody treatment and these signalling pathways in AOM/DSS mice tumours, we evaluated the expression of p-STAT3 and NF-κB p65 in the tumours of the MAdCAM- 1 Ab and IgG Ab groups. The percentage of nuclear staining for p-STAT3 and NF-κB p65 in the MAdCAM-1 Ab group was significantly lower than that in the IgG Ab group (Figure 7B, C).

**Figure 7.**
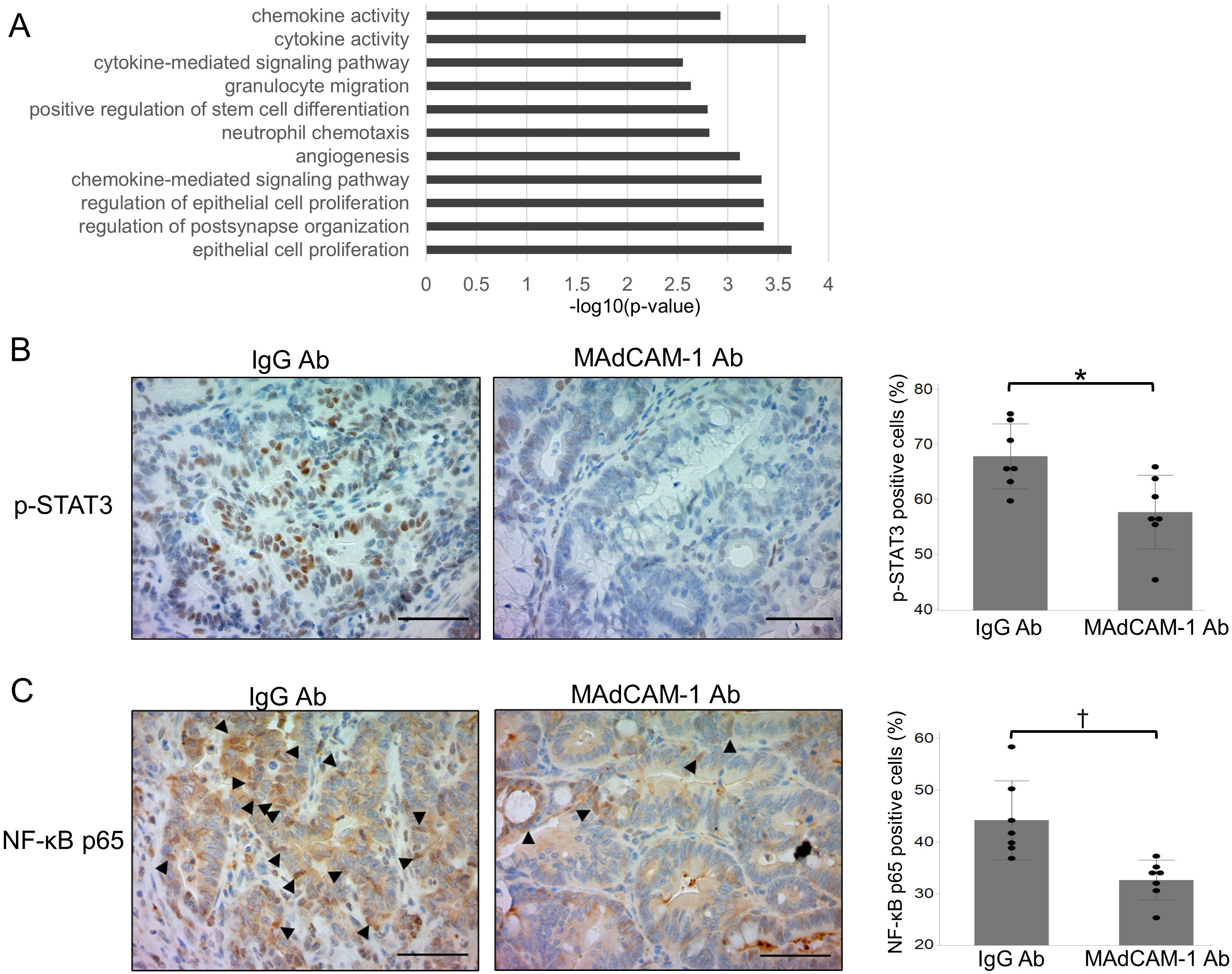
Inflammatory signal alteration by the MAdCAM-1 antibody treatment in colon tumours of the AOM/DSS mouse model. (A) Representative downregulated biological processes in the MAdCAM-1 Ab group compared to those in the IgG Ab group identified using gene ontology analyses. (B) Representative immunohistochemical images of p-STAT3, as a representative downstream of IL-6 signal, in colon tumours of the IgG Ab (n=7) and MAdCAM-1 Ab (n=7) groups. The percentage of p-STAT3-positive tumour cells was significantly higher in the IgG Ab group than in the MAdCAM-1Ab group. Scale bar, 50 μm. (Original magnification,×400). (C) Representative immunohistochemical staining images of nuclear NF-κB p65, as a representative downstream of TNF-α signal, in colon tumours of the IgG Ab (n=7) and MAdCAM-1 Ab (n=7) groups. The percentage of nuclear NF-κB p65-positive tumour cells was significantly higher in the IgG Ab group than in the MAdCAM-1Ab group. The data are expressed as the mean ± SD. *P<0.05, †P<0.01. Scale bar, 50 μm. (Original magnification, ×400). MAdCAM-1, mucosal addressin cell adhesion molecule-1.

## Discussion

Our clinical data demonstrated that the numbers of MAdCAM-1-positive vessels and infiltrating CD8-, CD68-, and CD163-positive immune cells were significantly higher in dysplasia/colitic cancer than in normal mucosa, SCRC, and UC mucosa. Moreover, MAdCAM-1-positive vessels in dysplasia/colitic cancer are associated with the infiltration of CD8- and CD163-positive immune cells. The AOM/DSS colitic cancer mouse model showed that tumour number, mean tumour diameter, tumour depth, Ki67 positivity, the number of CD8-, CD68-, CD163-positive immune cells, and the expression levels of inflammatory cytokines (IL-6 and TNFα) and tumour-promoting transcription factors (p-STAT3 and NF-κB p65) were significantly lower in the MAdCAM-1 Ab group than in the control IgG Ab group.

MAdCAM-1 is expressed in the endothelial cells of the intestinal mucosa and gut-associated lymphoid tissue. The interaction of MAdCAM-1-positive endothelial cells and intestinal-specific immune cells expressing α4β7 integrin promote the upregulation of immune cell migration to the intestinal mucosa (7). The increased MAdCAM-1- positive vessels in UC mucosa causes colitis by enhancing the migration of immune cells into the intestinal tract (13). In colorectal carcinogenesis, several inflammatory cytokines secreted by migrating immune cells have been reported to cause tumour initiation and progression via the inflammation/DNA damage axis (14), (15). Das et al. reported that the lack of β7 integrin, which binds specifically to MAdCAM-1, impairs tumour development (16). This study showed that the number of MAdCAM-1-positive vessels was higher in dysplasia/colitic cancers than in UC. Furthermore, the number of MAdCAM-1-positive vessels correlated with infiltration levels of CD163-positive macrophages and CD8-positive T lymphocytes in patients with dysplasia/colitic cancer. Interestingly, Zeissig et al. reported that treatment with vedolizumab, an α4β7 integrin inhibitor, reduced T cell and macrophage levels in the gastrointestinal tract (17). These observations suggest the importance of the interaction between MAdCAM-1-positive endothelial cells/integrin-positive immune cell axis in the dysplasia/carcinoma sequence of inflammatory UC mucosa.

Anti-inflammatory drugs, such as TNFα inhibitors and 5-amino salicylates, may inhibit tumour initiation and progression in colitic cancer (18, 19). In contrast, a representative anti-inflammatory steroid glucocorticoid promoted colitic cancer progression in AOM/DSS mouse model via suppression of immune cell infiltration into UC mucosa (20, 21). Therefore, the suppressive significance of immune cell infiltration into the colorectal mucosa in carcinogenesis remains controversial. MAdCAM-1 blockade suppresses inflammatory immune cell migration into the UC mucosa (17, 22). However, it is unclear whether this suppression inhibits colitic cancer progression or not (6). In this study, we showed that MAdCAM-1 blockade in a mouse model of colitic cancer resulted in a decreased inflammatory score, tumour number, tumour size, tumour depth, and Ki67 positivity. These data suggest that anti-inflammatory treatment with MAdCAM-1 blockade might have an inhibitory effect on the progression of colitic cancer in UC, unlike steroid treatment.

In clinical trials with patients with IBD, inhibition of the α4β7 integrin-MAdCAM-1 axis reduced not only the infiltration of B and T cells but also that of macrophages into the gastrointestinal tract, resulting in the amelioration of gastroenteritis (4). Furthermore, in an experimental colitis mouse model, the administration of MAdCAM-1 antibody improved colitis by reducing the infiltration of B cells, T cells, and macrophages into the colon mucosa, consistent with clinical trials, and ameliorated colitis (17, 23). However, few studies have addressed the therapeutic significance and molecular mechanism of MAdCAM-1 antibody treatment in colitic cancer carcinogenesis in UC patients and colitis mouse models. In this study, we demonstrated that MAdCAM-1 antibody treatment significantly reduced the number of tumours, tumour size, infiltration of macrophages and CD8-positive T cells, and tumoral expression of TNFα, IL-6, NF-kB, and p-STAT3. TNFα and IL6 are mainly secreted from macrophages, and the activation of the TNFα/NF-κB and IL-6/STAT3 pathways has contributed to the invasion and proliferation of several cancers, including colitic cancer (3, 24, 25). These data suggest that MAdCAM-1 blockage suppresses colitic cancer severity and carcinogenesis by regulating macrophage-associated inflammatory signals, such as TNFα/NF-kB and IL-6/STAT3.

In conclusion, this study showed that dysplasia/colitic cancer specimens exhibited increased MAdCAM-1-positive vessels with infiltration of CD8-, CD68-, and CD163- positive immune cells compared to SCRC specimens. Furthermore, MAdCAM-1 inhibitory antibody suppressed infiltration of CD8-, CD68-, and CD163-positive immune cells into the inflammatory intestinal mucosa; IL6/p-STAT3 and TNFα/NF-κB pathways; colon carcinogenesis; and tumour aggressiveness in AOM/DSS mice. Thus, a MAdCAM-1 targeting strategy, a therapeutic candidate against UC, could potentially control UC severity, carcinogenesis, and tumour progression by regulating inflammation/immune cell infiltration in patients with UC.

## Methods

### Patients and samples

Eleven patients (eight male and three female patients) with UC who underwent surgical resection at Gunma University Hospital and Maebashi Red Cross Hospital between 1999 and 2018 were included in this study. The median age of the patients was 54 years (range 37–76 years). One patient had dysplasia alone. Two patients had two or more tumours, and all high-grade dysplastic and cancerous lesions were evaluated; patients with low-grade dysplasia were not included in the study. Seventeen patients with SCRC (12 male and five female patients) who underwent partial colectomy at Gunma University Hospital between 1999 and 2010 were randomly selected and included in this study. Supplemental Table 3 summarises the patient information. For an accurate pathological diagnosis of dysplastic and cancerous lesions in patients with UC, all sections were evaluated by a specialised pathologist, Dr. Yao T (Department of Human Pathology, Juntendo University Graduate School of Medicine).

### Immunohistochemical staining

Paraffin-embedded blocks were cut into 4 µm-thick sections and mounted on glass slides. Sections were deparaffinised in xylene and dehydrated in alcohol. Endogenous peroxidase activity was inhibited by incubation with 0.3% H_2_O_2_/methanol for 30 min at room temperature. After rehydration through a graded series of ethanol treatments, antigen retrieval was performed using an Immunosaver (Nisshin EM, Tokyo, Japan) at 98–100 °C for 45 min. Nonspecific binding sites were blocked by incubation with Protein Block Serum-Free (Dako, Carpinteria, CA, USA) for 30 min. Next, the sections from the clinical samples were incubated overnight at 4 °C with primary antibodies against MAdCAM-1 (Proteintech, Chicago, IL, USA, 2A12E8, mouse mAb, 1:500 dilution); CD31, (Abcam, Cambridge, MA,

USA, ab28364, rabbit mAb, 1:100 dilution), a vascular endothelium maker (26); CD8 (DAKO, C8/144B, mouse mAb, 1:100 dilution), a cytotoxic T cell marker; CD68 (Abcam, ab955, mouse mAb, 1:100 dilution), a pan-macrophage marker; CD163 (Cell Signalling Technology, Beverly, MA, USA, D6U1J, rabbit mAb, 1:500 dilution), a tumour-associated macrophage marker; and FOXP3 (Abcam, ab22510, mouse mAb, 1:100 dilution), a regulatory T cell marker. The sections from mice were incubated overnight at 4 °C with primary antibodies against CD8 (Abcam, EPR20305, rabbit mAb, 1:1000 dilution), CD68 (Abcam, ab955, mouse mAb, 1:1000 dilution), CD163 (Abcam, EPR19518, rabbit mAb, 1:500 dilution), FOXP3 (Cell Signaling Technology, D6O8R, rabbit mAb, 1:200 dilution), Ki67 (Cell Signaling Technology, D3B5, rabbit mAb, 1:200 dilution) as a cell proliferation marker, p-STAT3 (Cell Signaling Technology, Tyr705 D3A7, rabbit mAb, 1:200 dilution), and NF-κB (Cell Signaling Technology, D14E12, rabbit mAb, 1:400 dilution). The Histofine Simple Stain MAX-PO (Multi) Kit (Nichirei, Tokyo, Japan) for the sections from clinical samples and Histofine Simple Stain mouse MAX-PO (R) Kit (Nichirei, Tokyo, Japan) for the sections from mouse tissues were used as the secondary antibody at room temperature for 30 min.

Chromogen 3,3’-diaminobenzidine tetrahydrochloride was applied as a 0.02% solution containing 0.005% H_2_O_2_ in 50 mM ammonium acetate-citrate acid buffer (pH 6.0). Nuclear counterstaining was performed using Mayer’s haematoxylin solution. Negative controls for immunohistochemical staining involved replacing the primary antibodies with phosphate-buffered saline in 0.1% bovine serum albumin and confirming a lack of staining.

### Immunohistochemical evaluation

We counted the number of MAdCAM-1-positive vessels in tissues from the patients and averaged the number in three fields of view at a magnification of ×200. The number of CD31-positive vessels was counted and used as the total number of vessels in the corresponding areas. The results in Figure 1C are expressed as the ratios of MAdCAM-1-positive vessels to CD31-positive vessels (8). In tissues from patients and AOM/DSS mice, the number of infiltrating CD8-, CD68-, CD163-, and FOXP3-positive cells per five high-powered fields (HPF, magnification ×400) was counted per each slide (27). The numbers of tumour cells positive for p-STAT3, Ki67, and NF-κB were determined by counting stained nuclei, and the positive rates were calculated by dividing the number of positive tumour cells by the total number of tumour cells per five HPFs (28).

### Murine colitis-associated tumorigenesis models (AOM/DSS model)

Fourteen female C57BL/6 mice aged 8 weeks were purchased from CLEA Japan Inc. (Tokyo, Japan). A colitic cancer mouse model was established via intraperitoneal injection (i.p) of 12.5 mg/kg AOM (Sigma-Aldrich, St. Louis, MO, USA) and by oral administration of 2.5% dextran sodium sulphate (DSS) (MP Biomedicals, Santa Ana, CA, USA). Eight-week-old female C57BL/6 mice were administered a single i.p. injection of AOM (12.5 mg/kg body weight). Seven days later, 2.5% DSS was administered in drinking water for seven days, followed by 14 days of regular water. Three cycles of DSS treatment were repeated (29). Anti-MAdCAM-1 antibody (MECA 367; Bio X Cell, Lebanon, PA, USA) or IgG antibody (2A3; Bio X Cell) at a dose of 100 μg/mouse was administered i.p. three times per week for each DSS cycle until one week after the end of DSS (22, 30, 31.

### Treatment and evaluation of experimental mouse

AOM/DSS mice were divided into two groups, with seven mice per group. The mice were monitored thrice weekly for body weight, stool consistency, and stool bleeding. Colitis severity was scored using the DAI (32), which was determined as follows: change in body weight loss (no weight loss or weight gain=0, 1–5% weight loss=1, 5– 10% weight loss=2, 10–15% weight loss=3, and >15% weight loss=4), stool consistency (normal and well-formed=0, very soft and unformed=2, and watery stool=4), and stool bleeding (normal colour stool=0, reddish colour stool=2, and bloody stool=4). The DAI score was calculated as the total of these scores and ranged from 0 (healthy) to 12 (severe colitis). Whole colon samples from all the mice were harvested on day 91. The colon was cut longitudinally along its main axis. Subsequently, we counted the number of tumours and measured the tumour size (average diameter in millimetres) in the harvested colon using a calliper. A tumour tissue section was stored in liquid nitrogen for quantitative real-time RT-PCR. Colon tissues were fixed in 10% buffered formalin and paraffin-embedded using the Swiss roll technique (33). For the histological evaluation of inflammation, paraffin-embedded colon sections were stained with H&E, and the sections were observed and evaluated by two researchers. The histological colitis score was determined using the following measures (34): goblet cell depletion (0, absent; 1, 1–9%; 2, 10–50%; 3, 51–100%), degree of inflammatory cell infiltration (0, absent; 1, increased presence of inflammatory cells; 2, infiltrates present also in the submucosa; 3, transmural), and crypt abscess (0, absent; 3, present). The histological colitis score was calculated as the total of these scores and ranged from 0 (healthy) to 9 (severe colitis).

### RT-PCR

Total RNA was extracted from tissues of AOM/DSS mouse tumours using the RNeasy Kit (Qiagen, Hilden, Germany), and the quantity of total RNA was measured using an ND-1000 spectrophotometer (NanoDrop Technologies, Wilmington, DE, USA). Quantitative real-time RT-PCR was performed using the GoTaq 1- Step RT-qPCR System (Promega, Madison, WI, USA) with a total reaction volume of 20 μL. The programme included four stages: reverse transcription at 37 °C for 15 min; RT inactivation and hot-start activation at 95 °C for 10 min; qPCR, 40 cycles of 95 °C for 10 s, 60 °C for 30 s, and 72 °C for 30 s; and dissociation at 60–95 °C (35). The relative levels of candidate genes were calculated using the 2^-ΔΔCT^ method. All primer sequences used in this study are listed in Supplemental Table 2. *GAPDH* was used to normalise the RNA input for all RT-PCR analyses.

### CAGE in colitic tumours of AOM/DSS mice

The total RNA of tumours in AOM/DSS mice was analysed using CAGE. CAGE library preparation, sequencing, mapping, gene expression analysis, motif discovery analysis, and GO enrichment analysis were performed using DNAFORM (Yokohama, Kanagawa, Japan). Total RNA quality was assessed using a Bioanalyzer (Agilent, Santa Clara, CA, USA) to ensure that the RNA integrity number was > 7.0. cDNAs were synthesised from total RNA using random primers. Ribose diols in the 5′ cap structures of RNA were oxidised and biotinylated. Biotinylated RNA/cDNA was selected using streptavidin beads (cap-trapping). After RNA digestion by RNaseONE/H and adaptor ligation to both ends of cDNA, double-stranded cDNA libraries (CAGE libraries) were constructed. The CAGE libraries were sequenced using single-end reads of 75nt on a NextSeq 500 instrument (Illumina, San Diego, CA, USA). The reads obtained (CAGE tags) were mapped to the mouse GRCm39 genome using BWA (version 0.7.17). The unmapped reads were mapped using HISAT2 (version 2.0.5). CAGE tag clustering, detection of differentially expressed genes, and motif discovery were performed using the pipeline RECLU (Ohmiya et al., BMC Genetics 2014, 15:269). The tag count data were clustered using the modified Paraclu programme. Clusters with count per million < 0.1 were discarded. Regions with 90% overlap between replicates were extracted using BED tools (version 2.12.0). Clusters with an irreproducible discovery rate ≥ 0.1 and clusters longer than 200 bp were discarded. Differentially expressed genes were detected using the edgeR package (version 3.22.5). For motif analysis, the genomic DNA sequences of the region from 200 bp upstream to 50 bp downstream of differentially expressed CAGE peaks were subjected to the de novo motif discovery tools AMD, GLAM2, DREME, and Weeder. The occurrence of these motifs was examined using FIMO. The similarity of consensus motifs and motifs in the JASPAR CORE 2016 vertebrates was evaluated using Tomtom. The list of differentially expressed genes detected by RECLU with a false discovery rate ≤ 0.05 were used for GO enrichment analysis using the cluster Profiler package (36).

### Statistics

All data are presented as mean ± SD. The JMP Pro 14.0 software package (SAS Institute Inc., Cary, NC, USA) was used to perform all statistical analyses. Statistical analysis was performed using Student’s t-test, Fisher’s exact test, and analysis of variance followed by Tukey’s test. Differences were considered statistically significant at P<0.05.

### Study approval

This study conformed to the tenets of the Declaration of Helsinki and was approved by the Institutional Review Board for Clinical Research at Gunma University Hospital, Maebashi, Gunma, Japan (approval number: HS2018-092). Patient consent was obtained by using the opt-out method. All mouse experiments were approved by the Gunma University Animal Care Committee and performed in compliance with the guidelines of the Institute for Laboratory Animal Research at Gunma University, Maebashi, Japan (approval number: 20-063).

## Author contributions

N. O., T. Yokobori, K. O., K. S., and H. S.: conceptualisation; N. O., T. Yokobori, K. O., K. S., and H. S.: funding; N. O., K. O., Y. S., T. M., M. I., and T. Oyama: resources; N. O.: methodology; N. O., E. O. B., H. Okami, Y. S., T. Yao, and T. Oyama: investigation; N. O. and T. Yokobori: writing - original draft; N. O., T. Yokobori, K. O., E. O. B., H. Okami, Y. S., T. S., T. Okada, A. S., M. Sakai, M. Sohda, T. M., M. I., H. Ogawa, T. Yao, T. Oyama, K. S., and H. S.: writing – review and editing; T. Yokobori: project administration; K. S. and H. S.: supervision. All authors read and approved the final version of the manuscript.

## Data Availability

All data produced in the present study are available upon reasonable request to the authors

## Acknowledgements

This study was supported by the Japanese Society for the Promotion of Science Grants-in-Aid for Scientific Research (KAKENNHI; 22K16528). The authors would like to thank MS. Yukiko Suto (Laboratory for Analytical Instruments, Education and Research Support Center, Graduate School of Medicine Gunma University) for their excellent technical assistance. This manuscript has been edited by English editing company, Editage, which was supported by Department of General Surgical Science, Graduate School of Medicine, Gunma University.

## Consent for publication

All authors agree to publish the paper in its present form.

## Notes

**Conflict of interest** The authors have declared that no conflict of interest exists.

### Competing Interest Statement

The authors have declared no competing interest.

### Author Declarations

This study conformed to the tenets of the Declaration of Helsinki and was approved by the Institutional Review Board for Clinical Research at Gunma University Hospital, Maebashi, Gunma, Japan (approval number: HS2018-092)

