## Supplementary figure for "Targeting MAdCAM-1 can prevent colitic cancer progression by suppressing immune cell infiltration and inflammatory signals"

**Supplemental Material**

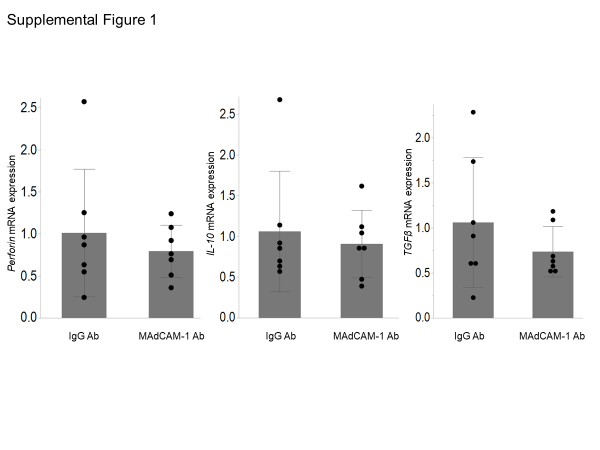

**Supplemental Figure 1. The expression of immune-related mRNA in colon tumours of the AOM/DSS mouse model.**

The relative expression levels of immune-related mRNA (perforin, *IL-10*, and *TGF-β*) were examined in the tumours of the IgG Ab (n = 7) and MAdCAM-1 Ab (n = 7) groups. *GAPDH* was used as the internal reference for each group. Data are expressed as the mean ± SD.

 MAdCAM-1, mucosal addressin cell adhesion molecule-1.

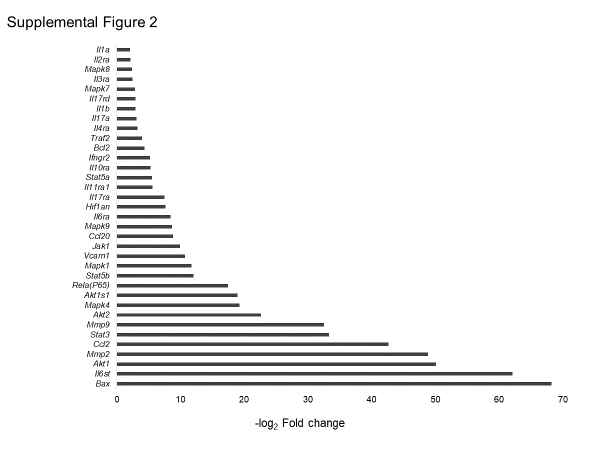

**Supplemental Figure 2. MAdCAM-1 antibody inhibits inflammatory and proliferation signalling in AOM/DSS mouse model.**

The fold change in mRNA levels of interleukin, STAT, and NF-κB-related genes was lower in the MAdCAM-1 Ab group than in the IgG Ab group.

STAT, signal transducer and activator of transcription; NF-κB, nuclear factor-κB; MAdCAM-1, mucosal addressin cell adhesion molecule-1.
